# The gender differences in the association between fear of pain and dental anxiety using the Japanese version of the Fear of Pain questionnaire-III: a cross-sectional study

**DOI:** 10.1101/2023.04.04.23288112

**Authors:** Mika Ogawa, Teppei Sago, Hirokazu Furukawa, Akihiro Saito

## Abstract

**Background:** The fear of pain is closely linked with chronic pain and the resultant impairment of daily life. It has been reported to have a correlation with dental anxiety, making its assessment crucial in dental practice. The present study aimed to develop a Japanese version of the Fear of Pain Questionnaire-III (FPQ-III), an international rating scale, to evaluate psychological characteristics and investigate its association with dental anxiety and gender differences.

**Methods:** After forward and backward translation and review, the Japanese version of the FPQ-III was administered to 400 internet monitors, and 100 of them were re-evaluated after a month. Convergent validity was assessed in relation to catastrophic thoughts of dental anxiety and pain, while discriminant validity was evaluated concerning the correlations between anxiety and depression. Confirmatory factor analysis was used to examine the factorial validity of the FPQ-III and a shortened version of the FPQ-9. Item response theory was applied to estimate the discriminative power of each item and draw a test information curve. Structural equation modeling was used to investigate the relationship between pain anxiety and dental anxiety and the gender differences in the model.

**Results:** Data from 400 participants (200 women [50.0%, mean 44.9 ± 14.5 years]) were analyzed. Total scores on the FPQ-III showed good internal validity, intra-examiner reliability, and discriminant validity, indicating convergent validity. Confirmatory factor analysis results supported a three-factor structure, and the FPQ-9 showed a good fit.

Discrimination was high, except for two items related to severe pain. Test information curves demonstrated that the FPQ-III and FPQ-9 were more accurate for latent characteristic values between -2 SD and +2 SD. Anxiety about medical pain fully mediated the relationship between fears of minor pain and dental anxiety. No gender differences were observed in this construct, and the factor means for anxiety about severe pain were significantly higher for women than for men.

**Conclusion:** The Japanese versions of the FPQ-III and FPQ-9 demonstrated high reliability and validity for measuring the Fear of Pain in this target population, with high accuracy for a wide range of latent characteristics. The fear of pain was deemed an endogenous factor that was linked to dental anxiety and common to both men and women.

## Introduction

Pain-related fear is a psychological factor that plays a role in the development and maintenance of chronic pain [1]. According to the gate-control theory, the central nervous system can influence pain perception [2]. As a biopsychosocial model of pain, chronic pain is considered to develop through a complex interaction of psychological, social, and biological factors [3]. In dentistry, pain-related fears contribute to maintaining oral-facial pain and are also associated with anxiety about dental treatment [4]. Therefore, assessing fear of pain is crucial in chronic pain research and dental clinical practice.

Self-report measures for pain-related fear and/or anxiety include the Pain Anxiety Symptom Scale [5] and the Fear of Pain Questionnaire-III (FPQ-III)[6]. The Pain Anxiety Symptom Scale assesses emotional, cognitive, and physiological changes during the pain experience, while the FPQ-III is a brief questionnaire that assesses fears of potentially painful experiences of various intensities that may occur in daily life.

The FPQ-III is a self-report questionnaire comprising 30 items that assess the intensity of fear of minor pain, severe pain, and pain associated with medical procedures. It has demonstrated high reliability and some construct validity [6]. It has shown high correlations with catastrophic thinking and fear of dental treatment but low correlations with general anxiety and depression, suggesting that these are distinct constructs[7-9]. There have been reports of a three-factor structure for factorial validity [6], a four-factor structure [10], a six-factor structure [11], and a three-factor structure assuming residual correlation [9]. Two shortened versions have been developed to date [10, 12]. Most studies on the FPQ-III have targeted the general population, with few studies in patients with chronic pain [12, 13]. One limitation of the reliability and validity of conventional methods is that they are entirely dependent on the characteristics of the target sample [12]. For example, results obtained in healthy individuals cannot be directly applied to a patient group. In contrast, item response theory (IRT) allows for the examination of the characteristics of the scale from the examinees’ ability values and the difficulty of the items, rather than being dependent on the sample [14].

Heritability has been reported for pain-related fears [15]. An association has been found between pain-related fears and dental anxiety, with the latest reports suggesting a common genetic basis for both. In the only genome-wide association study of the three subscales, fear of minor pain was reported to be the only phenotype significantly associated with the locus [16]. Dental anxiety is thought to have both external factors, such as negative dental treatment experiences and family learning of dental anxiety, and internal factors, such as genes and cognitive distortions [17, 18]. Based on the above, we hypothesize that anxiety about mild pain may be related to dental anxiety via anxiety about medical care. Furthermore, although pain anxiety is considered a type of internal factor for dental anxiety, there are no reports examining pain anxiety and external factors such as painful dental treatment experiences simultaneously.

The FPQ-III has been translated from English into four languages [8, 9, 11, 19], but a Japanese version has not yet been produced. Additionally, to our knowledge, there has been no scrutiny of the items using IRT. The first aim of this study was to develop a Japanese version of the FPQ-III and to examine its reliability and validity in a conventional manner and item response theory targeting the general Japanese population. The second aim was to examine whether medical-related pain mediates the relationship between anxiety about minor pain and dental anxiety; the third aim was to examine the pain-related fears and dental anxiety using structural equation modeling and to examine whether there are gender differences in this relationship. This study hypothesizes that pain related to medical treatment mediates the relationship between anxiety about minor pain and dental anxiety (hypothesis 1). Pain anxiety is associated with dental anxiety even after adjusting for the effect of negative dental care experiences (hypothesis 2).

## Methods

The study was approved by the Ethical Committee of Fukuoka Dental College (approval number: 586). The original author’s permission was obtained to develop a Japanese-language version. The FPQ-III was translated into Japanese using the back-translation method. Two fluent English speakers, one dentist, and one psychologist, separately translated the FPQ-III into English. Another dentist and a psychologist then merged these translations. An expert fluent in English, experienced in back-translation, retranslated the integrated Japanese version into English. Another translator, who had not been involved in the back-translation process, reviewed the original and back-translated versions. The internet survey was conducted in Japan in June 2022. All participants were internet monitors for MSS Inc. (Tokyo, Japan) who lived in Japan and were aged 20 or over. The target sample size was set at 400 based on previous studies [7, 9]. There were no exclusion criteria. A total of 10000 targets among the 307722 internet monitors received an e-mail invitation to participate in our survey from the research company. When the respondents visited the website for the survey, the policy for the use of data and the protection of personal information was displayed. Only those who agreed with the procedure were allowed to answer the questionnaire. The survey was closed when 440 participants had completed the questionnaire. MSS Inc. used its own protocols to clean the data and eliminate untruthful respondents. For reliability studies using the retest method, the FPQ-III was studied again one month later in 100 randomly selected participants, with a 50% male/female ratio.

### Measures

All participants were asked to provide details about sociodemographic characteristics (i.e., age, gender, occupation, and educational level) and clinical information (i.e., presence of chronic pain lasting more than three months, Location and frequency of chronic pain). They were also asked whether they had experienced painful dental treatment, painful medical treatment, broken bones, and road traffic accidents. Current pain was measured using the Numerical Rating Scale, which asked respondents to “Please answer the intensity of the pain you have felt most often during the past month, with no pain as 0 and worst pain as 10”.

Fear of pain was measured using the FPQ-III, a 30-item self-administered questionnaire that assesses fear of painful experiences that may be encountered in everyday life and healthcare settings [6]. Each item was obtained on a five-point Likert scale ranging from not at all to extreme, with an overall score ranging from 30 to 150. Three subscales (severe, minor, and medical pain) exist, with ten items in each. Good internal consistency, retest reliability, and convergent and discriminant validity are reported. There is a shortened 23-item version by Asmundson et al. [10] and a 9-item version by the original authors [12].

Anxiety and depression were assessed using the Japanese version of the Hospital Anxiety and Depression Scale, a self-administered scale [20]. It consists of seven items on anxiety and seven items on depression and is used for screening purposes in outpatient clinics and health check-ups due to its simplicity. The responses were obtained on a four-point scale. The range is 0-21 points each. The Japanese version has been reported to have high reliability and validity [21].

Dental anxiety was measured with the reliable and valid Japanese version of the Modified Dental Anxiety Scale (MDAS) [22], a 5-item questionnaire that assesses anxiety in five situations: going for treatment tomorrow, sitting in the waiting room, having one’s tooth drilled, having one’s teeth scaled and polished, and receiving a local anesthetic injection. Responses are recorded on a 5-point Likert-type scale ranging from “not anxious” to “extremely anxious” that sum up to a total score (range 5–25). Higher scores indicate greater dental anxiety. The two established factors of MDAS were calculated as anticipatory dental anxiety (items 1 and 2; range = 2–10) and treatment dental anxiety (items 3, 4, and 5; range = 3–15) [23, 24]. The Japanese version of MDAS has been reported to be one-factor [25, 26].

Pain Catastrophizing was measured using the Pain Catastrophizing Scale, a valid and reliable Japanese 13-item self-report measure was used to assess three components of catastrophizing: rumination, magnification, and helplessness [27, 28]. Responses were measured with a five-point Likert scale ranging from “strongly disagree” to “strongly agree.” Total scores range from 13 to 65.

### Statistical analyses

Sample demographics and mean values for the FPQ and other scales were calculated. Dental anxiety and catastrophic thoughts of pain were assessed using Pearson’s correlation coefficients to investigate convergent validity and anxiety and depression to investigate discriminant validity. Reliability was assessed using internal consistency and retest reliability methods. Cronbach’s assessed internal consistency of the total scale and subscales; values >.70 were considered acceptable [29]. The Intraclass Correlation Coefficient (ICC) was employed to evaluate test-retest reliability. As a rule of thumb, ICC values between .61 and .80 indicate moderate reliability, and those between .81 and .90 substantial reliability [30].

#### Item Response Theory

Item response theory was used to estimate the discriminative power of each item and draw a test information curve [14, 31]. Before the main IRT, categorical factors analysis and a preliminary IRT were conducted to remove items with a factor pattern of less than 0.35, a discriminative power of less than 0.2, and a difficulty level of more than 4.

#### Structural Equation Modeling

The factorial validity of the FPQ-III and the shortened version was examined using confirmatory factor analysis. The following fit indices were used: chi-square and its significance, the comparative fit index (CFI), the root mean square error of approximation (RMSEA), and standardized root mean square residual (SRMR). The values of X2/df < 5, CFI > 0.90, and RMSEA < 0.08 indicate a reasonably good fit [32]. The best model has the smallest Akaike information criterion (AIC) and Bayesian information criterion (BIC)[32].

#### Measurement invariance

To examine the equality of the covariance structure between men and women, we constructed models with the following constraints.

Configural Invariance Model: a model in which the observed variables measuring the nuclear factor are equal across populations

Metric Invariance Model: a model in which, across populations, the factor patterns measuring each factor are equal

Scalar Invariance Model: in addition to Metric Model, a model in which the intercepts measuring each is equal

Strict Invariance Model: in addition to Scalar Invariance Model, a model in which the residuals measuring each item are equal

Each model was compared to the previous model regarding changes in CFI (ΔCFI), RMSEA, AIC, and BIC employing the following cut-off values: .010 for ΔCFI [33]. SPSS Statistics software version 27 (IBM SPSS, Armonk, NY, USA) for description and factor analysis for exploration. IRT and SEM were conducted using R version 4.0.0 (R Foundation for Statistical Computing, Vienna, Austria) and Package “ltm” [34] and “lavaan” [35]. All tests were conducted at a significance level of 0.05.

## Results

Data from 400 subjects who passed the quality checks (200 women [50.0%, mean 44.9 ± 14.5 years]) were analyzed. Socio-demographics of the participants are shown in Table 1. About half of the respondents were company employees, and about half had a university degree. About 60% felt they had chronic pain, which was mild with an average NRS of 2.5.

**Table 1.**
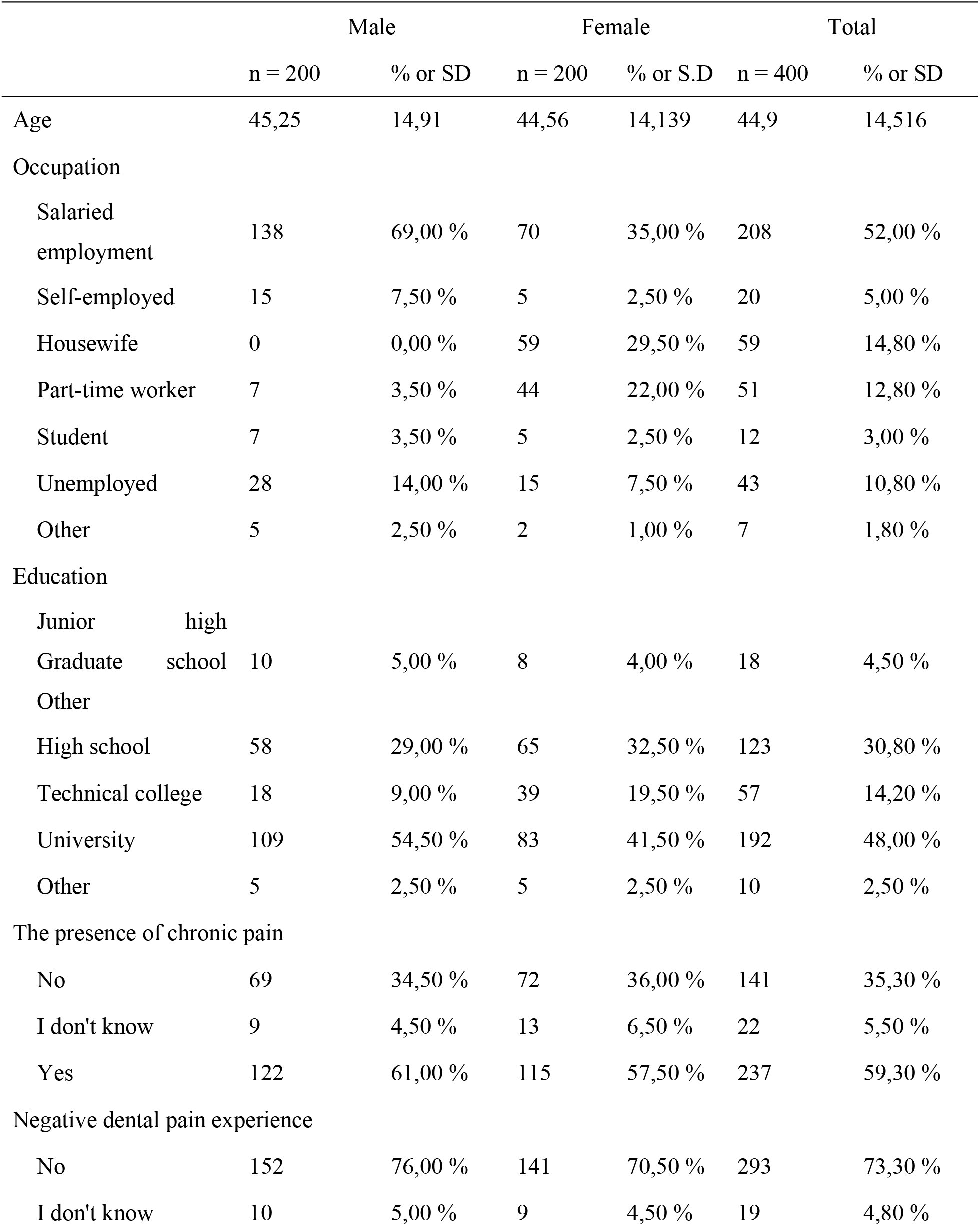
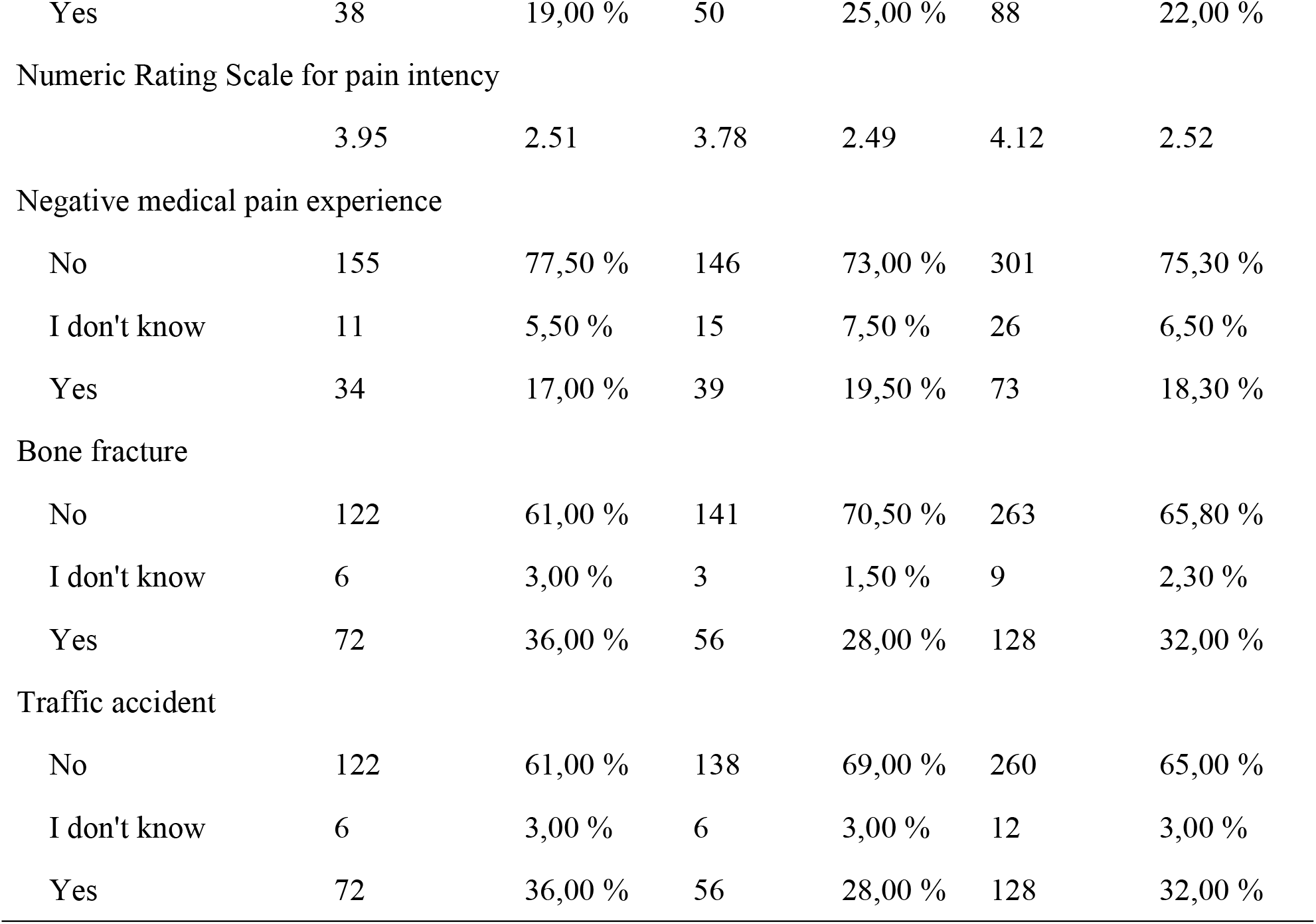
Sociodemographic variables and experiences of the participants.

The translated version of FPQ-III is shown in S1.

The means of the total FPQ-III scores and subscales and their gender differences are shown in Table 2. For all scores, females were significantly higher than males.

**Table 2.** Mean FPQ-III total scores and subscales and their gender differences.

|  | Total (n = 400) |  | Male (n = 200) |  | Female (n = 200) |  | P value |
| --- | --- | --- | --- | --- | --- | --- | --- |
|  | Mean | S.D. | Mean | S.D. | Mean | S.D. |  |
| Total | 95.66 | 22.92 | 91.78 | 22.67 | 99.54 | 22.55 | <0.001 |
| Severe | 39.35 | 7.63 | 37.44 | 7.65 | 41.25 | 7.13 | <0.001 |
| Minor | 26.96 | 9.07 | 26.02 | 8.98 | 27.89 | 9.07 | 0.04 |
| Medical | 29.36 | 9.11 | 28.32 | 8.90 | 30.40 | 9.22 | 0.02 |

Abbreviations: FPQ, Fear of Pain Questionnaire.

### Reliability and internal consistency

Total scores on the FPQ-III showed good internal validity and within-examiner reliability (Table 3).

**Table 3.** Internal consistency and test-retest reliability.

| | Cronbach $\alpha$ | ICC | ICC 95% CI | |
| --- | --- | --- | --- | --- |
| Severe | 0,894 | 0,676 | 0,555 | 0,770 |
| Minor | 0,918 | 0,823 | 0,749 | 0,878 |
| Medical | 0,918 | 0,854 | 0,791 | 0,899 |
| Total | 0,958 | 0,812 | 0,734 | 0,870 |

Abbreviations: ICC, Intraclass Correlation Coefficient.

### Validity

Table 4 shows FPQ-III and depressive tendency and anxiety levels showed no to weak positive correlation and had discriminant validity (r = -0.07, p = 0.16; r = 0.26, p < 0.01). On the other hand, moderate positive correlations were found between the FPQ-III and dental anxiety and catastrophic thinking, showing convergent validity (r = 0.52, p < 0.01; r = 0.49, p < 0.01).

**Table 4.** Pearson’s correlation for divergent and convergent validity.

|  | Anxiety | Depression | MDAS | PCS |
| --- | --- | --- | --- | --- |
| FPQ total | .263** | -0,071 | .517** | .492** |
| Severe | 0,087 | -.260** | .343** | .347** |
| Minor | .284** | 0,016 | .448** | .426** |
| Medical | .306** | 0,023 | .568** | .523** |

Abbreviations: FPQ, Fear of Pain Questionnaire; MDAS, Modified Dental Anxiety
Scale; PCS, Pain Catastrophizing Scale.

Confirmatory factor analysis showed a poor fit of the three-factor structure (FPQ-J model). However, the goodness of fit improved when error correlations were set for items related to injection, fracture, and dentistry as Di Tella et al. showed [9]; the FPQ-9 [12] showed a good fit (Table 5). The results of Di Tella’s model are presented in S2.

**Table 5.**
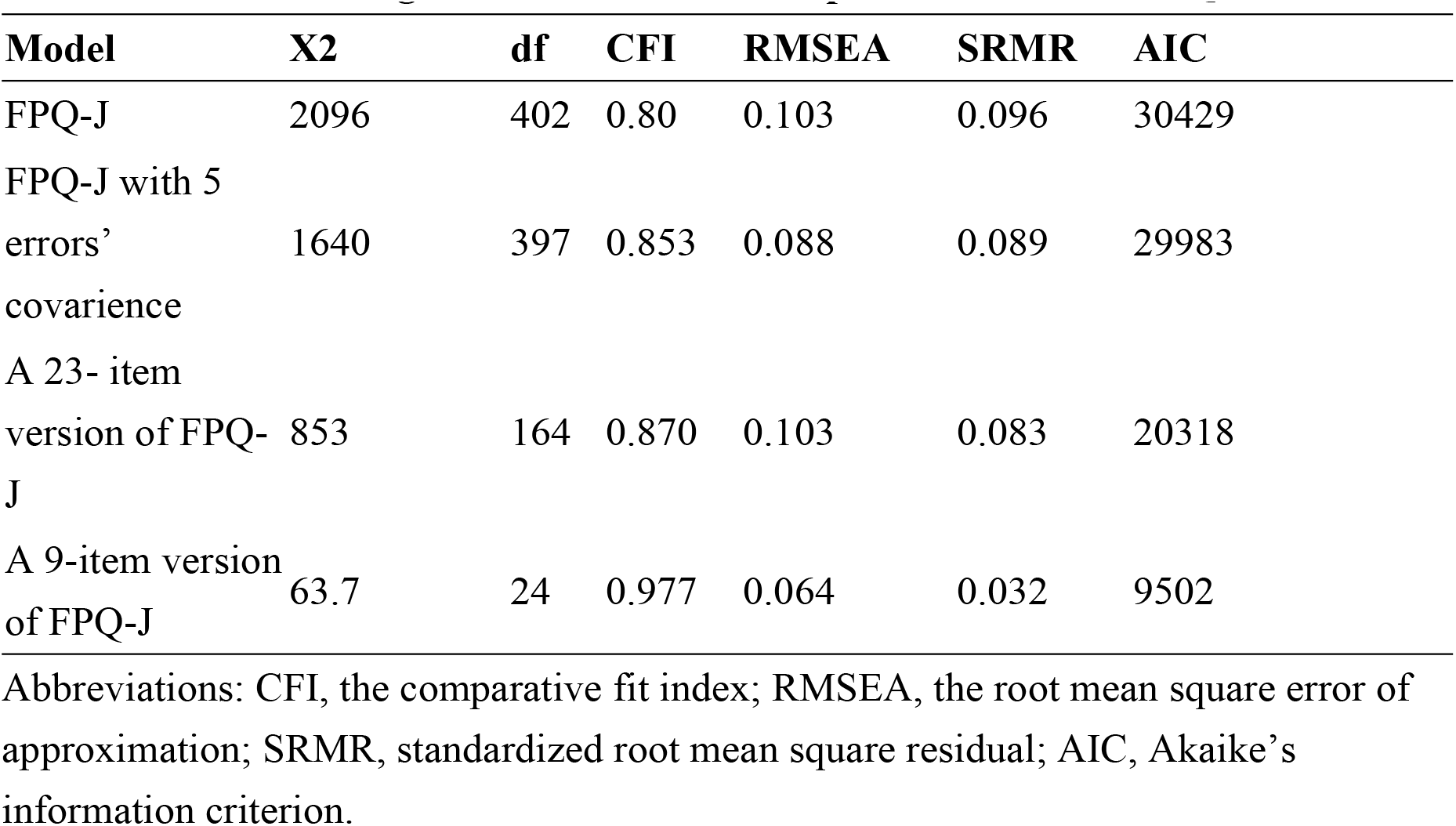
CFA model’s goodness of fit for the Japanese version of FPQ-III.

### Item analysis

Discrimination was high except for two items related to severe pain (Table 6), and test information curves showed that the FPQ-III and FPQ-9 were more accurate in targets with latent characteristic values between -2 SD and +2 SD (Fig 2).

**Table 6.**
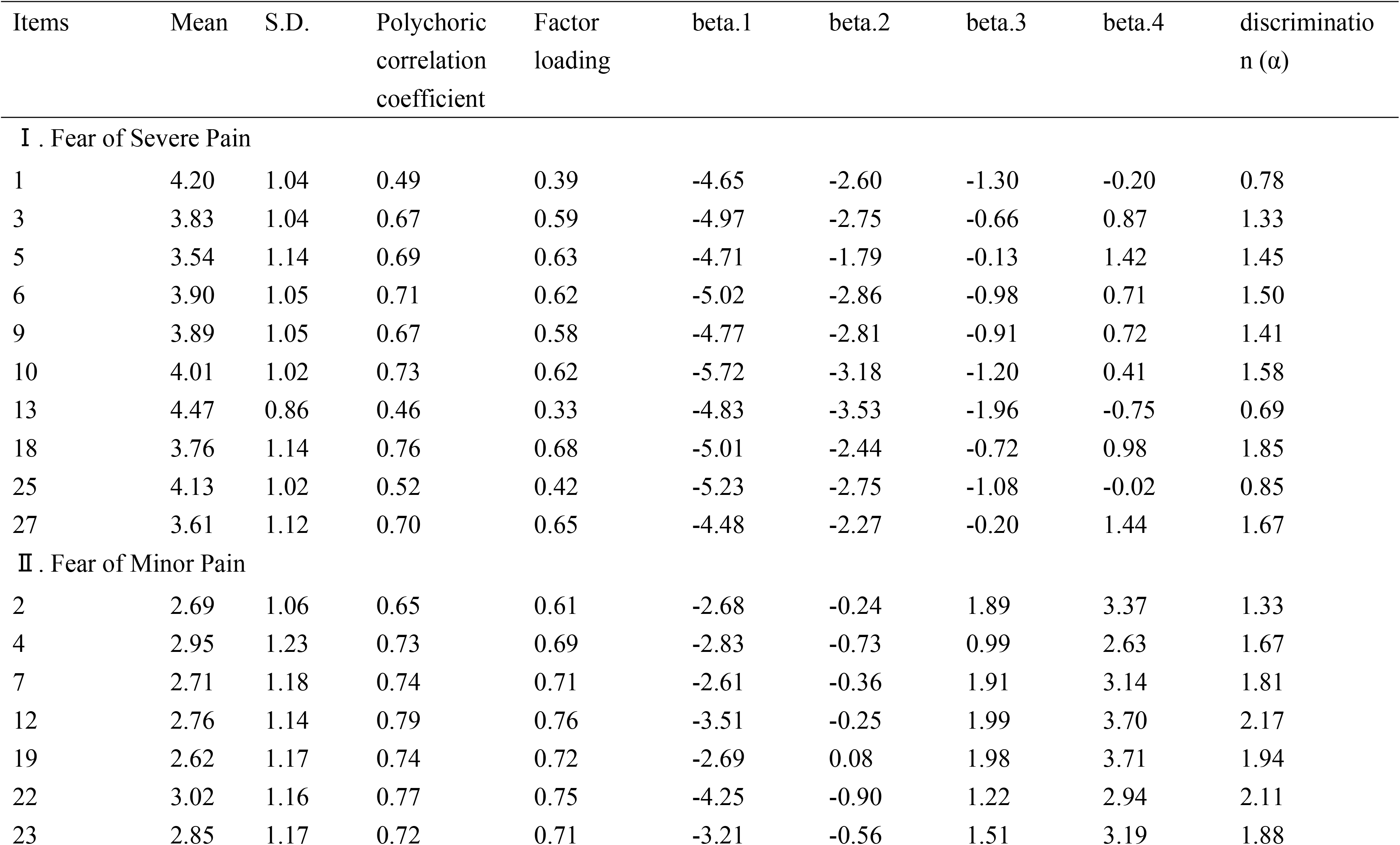
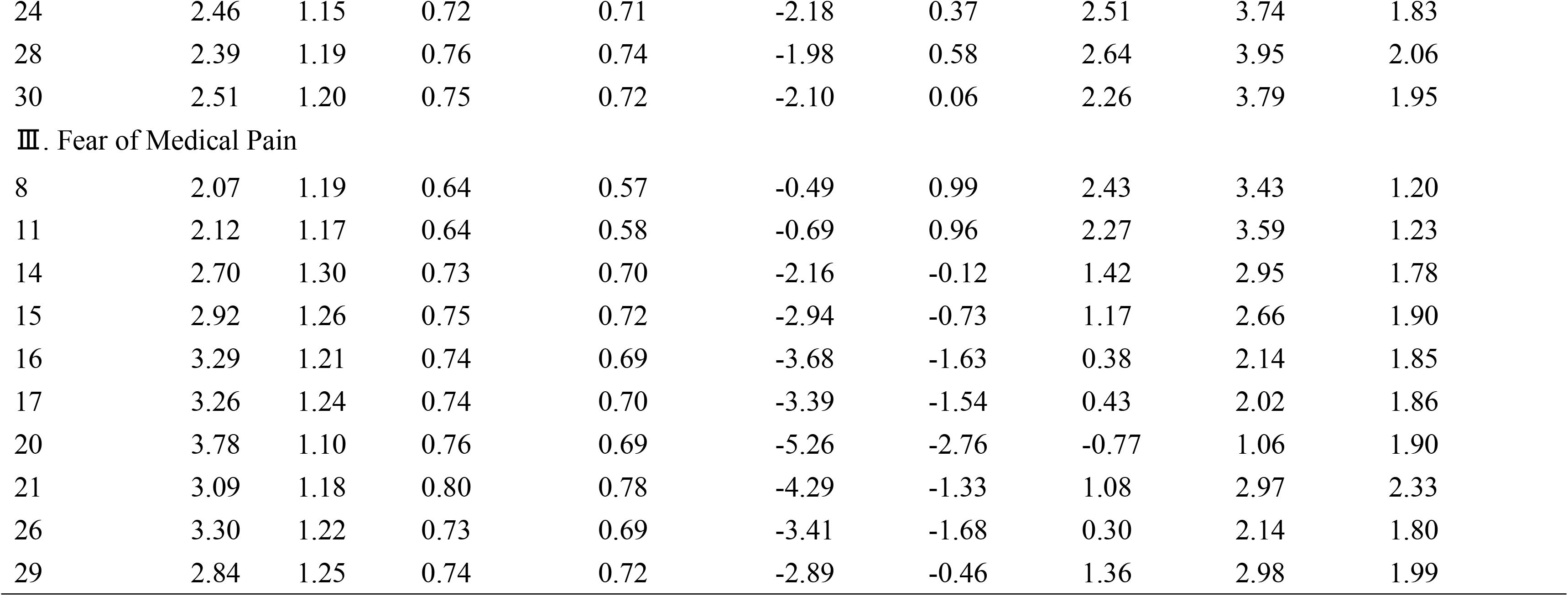
Results of item-specific descriptive statistics and item analysis.

**Fig 1.**
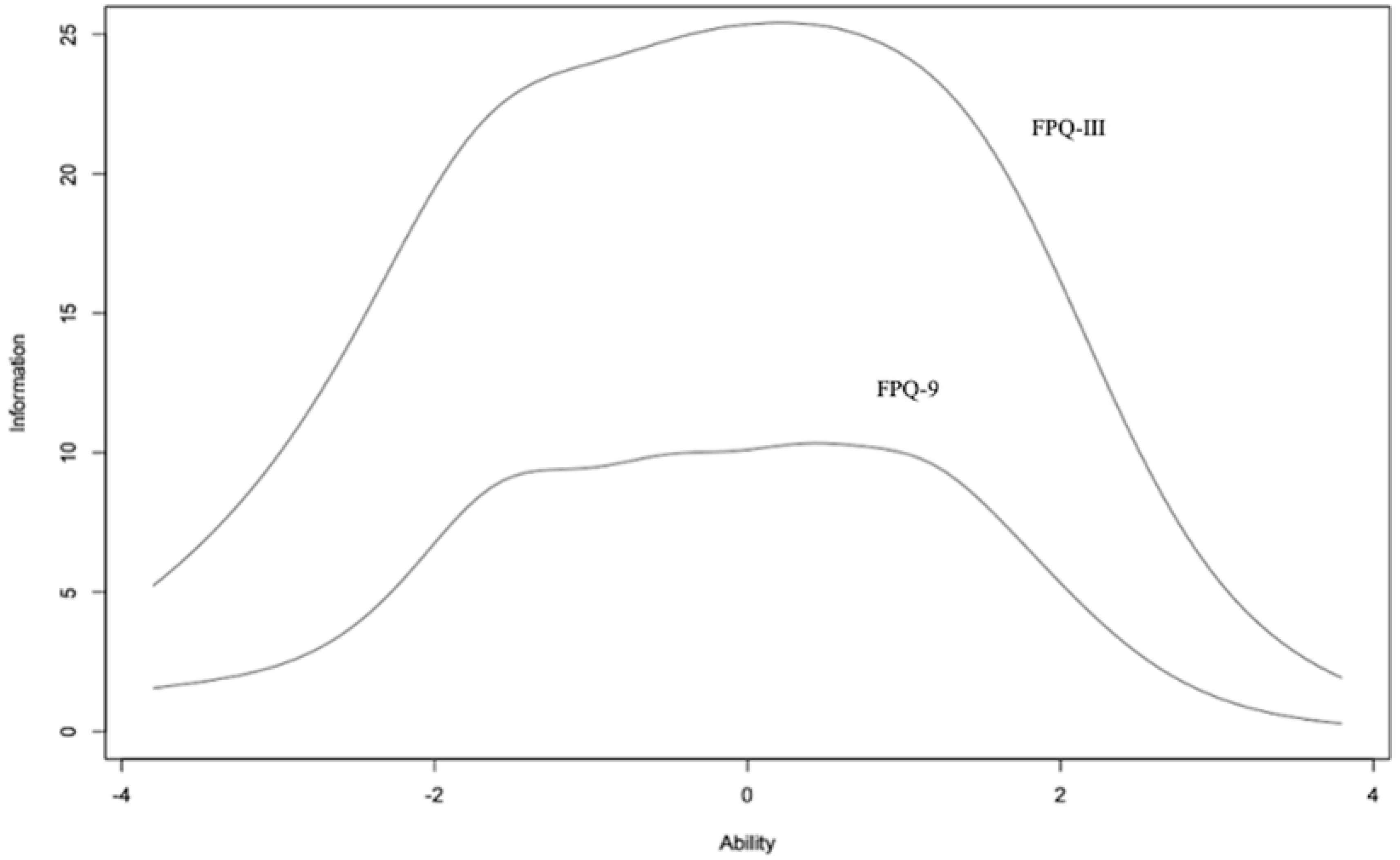
Test information functions for FPQ-III and a shortened version. FPQ-III: fear of pain questionnaire III; FPQ-9: nine items of fear of pain questionnaire III

**Fig 2.**
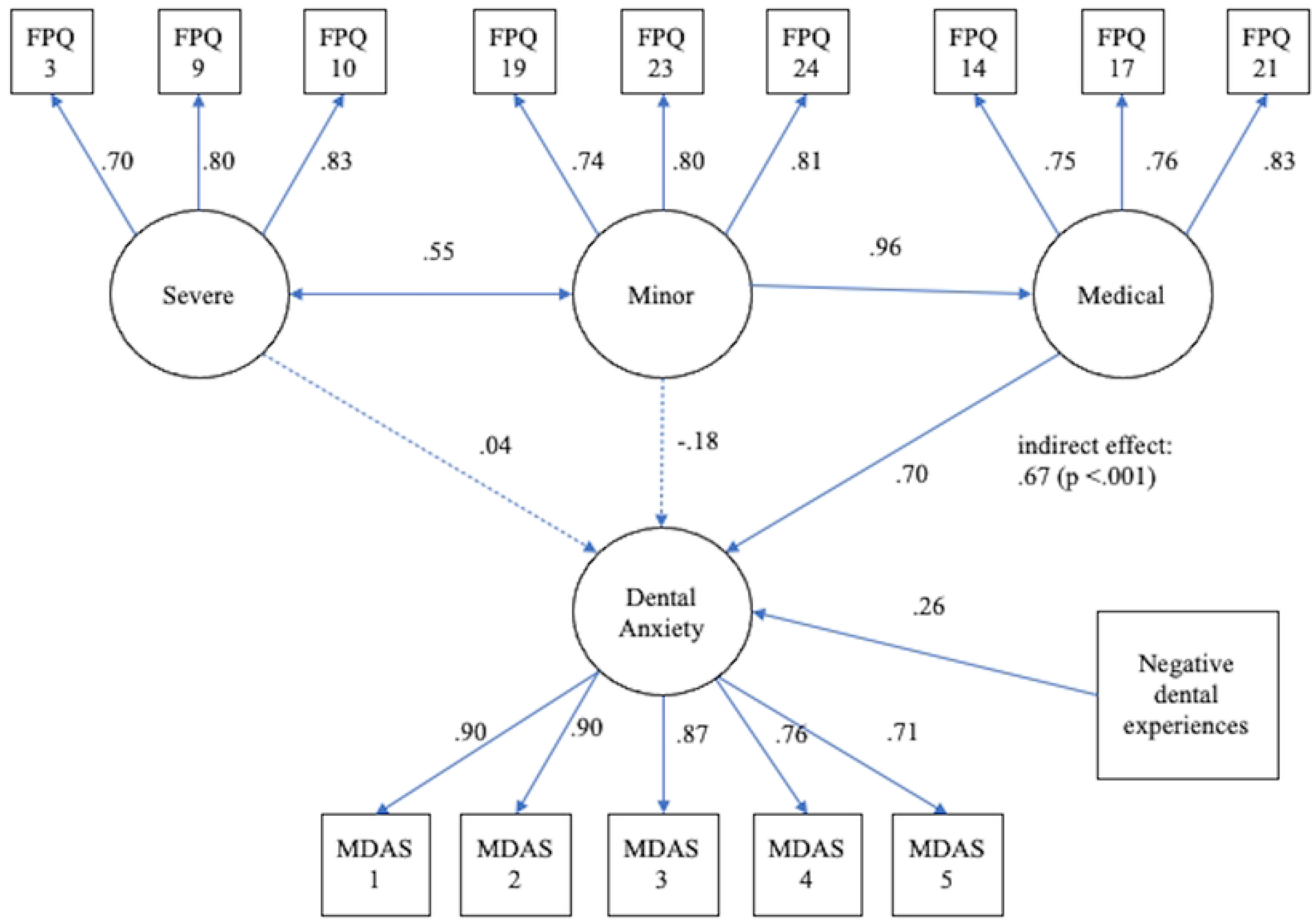
The estimated structural equation model with standardized coefficients for the interrelationships of dental anxiety, fear of severe/mild pain, and negative dental experiences. Dotted lines indicate non-significant paths

### Structural Equation Modeling (SEM)

Before performing SEM, the factor structure of the MDAS was checked: an EFA using SPSS showed that the decay of eigenvalues was 3.38, 0.49, and 0.37, supporting a one-factor structure according to the Guttman criterion and scree plot. The one-factor model was adopted because the Japanese version of the MDAS also showed a one-factor structure in previous studies [25, 26].

Fears of minor pain are related to dental anxiety through fears of medical pain-related fears. Furthermore, severe pain-related fears are related to dental anxiety in correlation with fears of minor pain and medical pain-related fears. A model was developed in which the experience of painful dental treatment is also associated with dental anxiety. The results are presented in Fig 2.

The direct effect of fears of minor pain on dental anxiety was not significant, with a significant indirect effect via fears about medical pain (indirect effect .672 95% confidence interval [.375-.968]). The results support hypothesis 1. Fears about severe pain was not significantly associated with dental anxiety. Furthermore, painful dental treatment experiences were significantly associated with dental anxiety. These results partially supported hypothesis 2.

A multi-group SEM was then used to examine whether there were gender differences in these results. The goodness of fit of the model is shown in Table 7. The invariant configuration model also showed no significant worsening. The introduction of path coefficients, fixed intercepts, and mean structure did not significantly deteriorate the goodness of fit. The strict model was therefore adopted. The mean structure was significantly higher for women than men for anxiety about severe pain and medical pain (.395, p < .001; .178. p = .02).

**Table 7.**
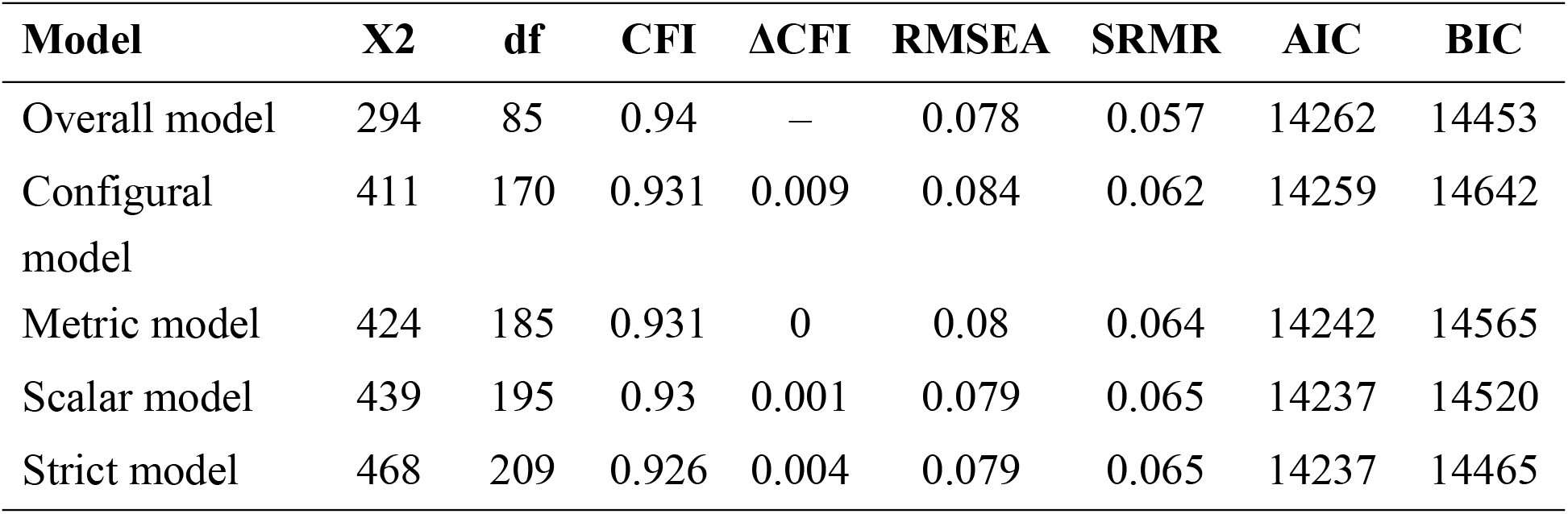
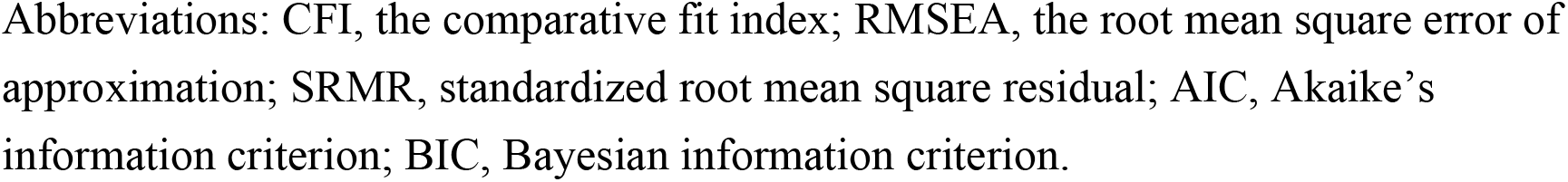
Invariances across gender.

| Model | X2 | df | CFI | ΔCFI | RMSEA | SRMR | AIC | BIC |
| --- | --- | --- | --- | --- | --- | --- | --- | --- |
| Overall model | 294 | 85 | 0.94 | – | 0.078 | 0.057 | 14262 | 14453 |
| Configural model | 411 | 170 | 0.931 | 0.009 | 0.084 | 0.062 | 14259 | 14642 |
| Metric model | 424 | 185 | 0.931 | 0 | 0.08 | 0.064 | 14242 | 14565 |
| Scalar model | 439 | 195 | 0.93 | 0.001 | 0.079 | 0.065 | 14237 | 14520 |
| Strict model | 468 | 209 | 0.926 | 0.004 | 0.079 | 0.065 | 14237 | 14465 |

Abbreviations: CFI, the comparative fit index; RMSEA, the root mean square error of approximation; SRMR, standardized root mean square residual; AIC, Akaike's information criterion; BIC, Bayesian information criterion.

## Discussion

The present study aimed to construct a Japanese version of the FPQ-III and examine its psychometric properties in a non-clinical Japanese sample. The three-factor structure, high reliability, and validity were found; IRT results showed good accuracy, particularly for the shortened 9-item version, to the same extent as the original version; SEM results showed that fears of minor pain were related to dental anxiety via medical pain-related fears, even after adjusting for the effect of negative dental treatment experiences. This structure did not differ between men and women.

The results of this study showed high internal consistency and retest reliability, as in other translated versions [6-9]. In line with previous studies, dental anxiety was moderately positively correlated with catastrophic thoughts about pain. Of the three subscales, only anxiety about severe pain showed a low correlation with depression, indicating heterogeneity.

This is the first study to analyze FPQ-III items using IRT. Anxiety about severe pain had a higher mean and a lower discrimination index than the other two subscales. In particular, 13: breaking your neck, 1: being in a car accident, and 25: being sick and feeling pain daily due to imminent death have high discrimination indices, indicating a ceiling effect. 9-item shortened versions remove these items and the test information curves also show high accuracy for participants with a wide range of latent characteristics. These suggest that the shortened 9-item version may be helpful.

The present study showed that fears of minor pain are related to dental anxiety via fears of pain-related fears of medical care, a construct common to both sexes. A previous GWAS-based study reported that of the three subscales of pain anxiety, only pain-related fears was a locus-related phenotype [16]. We suggested a mechanism by which differences in sensitivity to pain differ at the genetic level and evolve into anxiety about fears of minor medical pain, such as injection pain, leading to anxiety about dental treatment.

The study found no gender differences in the relationship between pain fears and dental anxiety. The mean values of latent factors were significantly higher in women only for severe and medical pain fears. This supports previous reports that fears about pain are higher in women than in men [9]. On the other hand, no differences were found in the factor means for dental anxiety between men and women, although a trend towards higher dental anxiety is also reported in women [22, 26].

The present study has several limitations. First, the participants in this study were from the general population, and the structure may differ, for example, in patients seen in pain clinics or those with strong dental anxiety. Secondly, the present study is an observational study design, so a causal relationship cannot be determined. However, the study has strengths in terms of sufficient sample size, a wide age range and an equitable gender ratio, and the use of IRT.

## Conclusion

In summary, the Japanese version of the FPQ-III was shown to be highly accurate for participants with a wide range of latent characteristics. Furthermore, fears of minor pain were suggested to be a common trait among men and women, leading to anxiety about medical treatment, particularly dental treatment.

## Data Availability

All relevant data are within the manuscript and its Supporting Information files.

## Supporting information

**S1 File. The Japanese version of the Fear of Pain Questionnaire-III**

**S1 Fig. Standardized solution of the FPQ-III three-factor model with five error items correlations**. MdP: Medical Pain; MP: Minor Pain; SP: Severe Pain

**S1 Dataset. This is raw data used in the present study**.

## Notes

### Competing Interest Statement

The authors have declared no competing interest.

### Funding Statement

Awardee: O.M. The name of the fund: JSPS KAKENHI: Grant-in-Aid for Early-Career ScientistsGrant Number: 20K18817. Funder: Japan Society for the Promotion of Science URL: https://www.jsps.go.jp/english/ The funder did not play any role in the study.

### Author Declarations

The study was approved by the Ethical Committee of Fukuoka Dental College (approval number: 586). Informed consent was not obtained for this survey as it was an anonymous research. An explanatory document was presented prior to the survey and only those who consented to the survey were transferred to the main survey.

